# Transmission characteristics of SARS-CoV-2 variants of concern Rapid Scoping Review

**DOI:** 10.1101/2021.04.23.21255515

**Authors:** Janet Curran, Justine Dol, Leah Boulos, Mari Somerville, Holly McCulloch, Marilyn MacDonald, Jason LeBlanc, Lisa Barrett, Todd Hatchette, Jeannette Comeau, Bearach Reynolds, Danielle Shin, Allyson Gallant, Helen Wong, Daniel Crowther, Ziwa Yu

## Abstract

**Background:** As of March 2021, three SARS-CoV-2 variants of concern (VOC) have been identified (B.1.1.7, B.1.351 and P.1) and been detected in over 111 countries. Despite their widespread circulation, little is known about their transmission characteristics. There is a need to understand current evidence on VOCs before practice and policy decisions can be made. This study aimed to map the evidence related to the transmission characteristics of three VOCs.

**Methods:** A rapid scoping review approach was used. Seven databases were searched on February 21, 2021 for terms related to VOCs, transmission, public health and health systems. A grey literature search was conducted on February 26, 2021. Title/abstracts were screened independently by one reviewer, while full texts were screened in duplicate. Data were extracted using a standardized form which was co-developed with infectious disease experts. A second data extractor verified the results. Studies were included if they reported on at least one of the VOCs and transmissibility. Animal studies and modeling studies were excluded. The final report was reviewed by content experts.

**Results:** Of the 1796 articles and 67 grey literature sources retrieved, 16 papers and 7 grey sources were included. Included studies used a wide range of designs and methods. The majority (n=20) reported on B.1.1.7. Risk of transmission, reported in 15 studies, was 45-71% higher for B.1.1.7 compared to non-VOCs, while R_0_ was 75-78% higher and the reported R_t_ ranged from 1.1-2.8. There was insufficient evidence on the transmission risk of B.1.35.1 and P.1. Twelve studies discussed the mechanism of transmission of VOCs. Evidence suggests an increase in viral load among VOCs based on cycle threshold values, and possible immune evasion due to increased ACE2 binding capacity of VOCs. However, findings should be interpreted with caution due to the variability in study designs and methods.

**Conclusion:** VOCs appear to be more transmissible than non-VOCs, however the mechanism of transmission is unclear. With majority of studies focusing on the B.1.1.7 VOC, more research is needed to build upon these preliminary findings. It is recommended that decision-makers continue to monitor VOCs and emerging evidence on this topic to inform public health policy.

## Introduction

COVID-19, caused by the virus SARS-CoV-2, was declared a worldwide pandemic on March 11^th^, 2020 by the World Health Organization (WHO).^1^ Since then, it has claimed over 2.45 million lives and 110 million cases have been identified as of February 19^th^, 2021.^2^ COVID-19 has had a significant impact worldwide, shutting down schools and workplaces, disrupting travel and trade, causing significant mortality and morbidity, particularly among vulnerable populations, and negatively impacting people’s mental health.^1,3^

A growing area of interest are Variants of Concern (VOCs). SARS-CoV-2 VOCs result in changes in transmissibility, clinical presentation and severity, or they may have an impact on countermeasures, including diagnostics, therapeutics and vaccines.^3^ As of February 20^th^, 2021, three VOCs have been identified from 3 different genetic lineages: B.1.1.7 (201/501.1.V1 or 202012/01) in the UK,^4^ B.1.351 (20H/501Y.V2) in South Africa,^5^ and P.1 (previously known as B.1.1.28.1) in Brazil.^6^ The first VOC identified was the B.1.1.7 on December 14^th^, 2020, which contained 23 nucleotide substitutions (including eight spike mutations among 14 non-synonymous mutations)^7^ and was not phylogenetically related to the SARS-CoV-2 virus.^3^ As of March 9, 2021, this variant was found in 111 countries/ territories/ areas. Similarly, the VOCs found in South Africa and Brazil have been detected in 58 and 32 countries, respectively.^8^

What remains unknown is the precise degree of impact these variants have on COVID-19 transmission, and the reason for the increased transmissibility, as well as the clinical impact of these strains on severe disease, hospitalization, and death. Previous studies have suggested that VOCs are more transmissible than the original strain, but there is limited evidence related to exactly how much more transmissible they are, due to the heterogeneity in study design and modeling approaches.^3^ There is a need to understand more about VOCs as changes to public health recommendations may be required to minimize the further spread of COVID-19.

In order to inform evidence-based recommendations that can reduce VOC-associated risk, health systems need to understand the existing literature and potential impacts of VOCs. Due to the emerging nature of this topic, no current or underway systematic reviews or scoping reviews on the topic were identified. Therefore, the aim of this rapid scoping review is to map the literature related to the transmission characteristics of the SARS-CoV-2 VOCs.

### Questions

This research brief presents evidence related to the following questions about transmission of the three major SARS-CoV-2 VOC as known in March 2021 (B.1.1.7 or UK variant; B.1.351 or South Africa variant; P.1 or Brazil variant):

1. How much more transmissible are they? (both in greater susceptibility to infection given exposure and in greater infectiousness once infected)?
2. Why are they more transmissible? (e.g., does the mechanism of transmission vary, can transmission happen with less exposure, and does the timing of peak viral load vary?)
3. What criteria can be used to define new variants of concern?

## Design

Rapid scoping review, following standardized rapid and scoping review guidelines.^9–11^

## Methods

An information specialist designed a broad, comprehensive search to retrieve all literature related to the VOCs. The electronic database search was executed on February 21^st^, 2021 in MEDLINE, Embase, the Cochrane Database of Systematic Reviews (CDSR) and Central Register of Controlled Trials (CENTRAL), Epistemonikos’ L·OVE on COVID-19, and medRxiv and bioRxiv concurrently. The electronic database search was followed up by a grey literature search, executed February 26^th^ to March 1^st^, 2021, in a list of specific websites in addition to broader searches of Google and Twitter. Only English-language searches were conducted, but non-English results were considered for inclusion. Full search details are shared in Appendix A.

Evidence specific to VOC transmission was identified and tagged during the screening process. For the purpose of this review, measures of VOC transmission included any of the following: decreases in PCR cycle threshold (Ct) values (as a proxy for viral load), changes in reproduction number, growth rate increases in specific lineages based on genomic data, markers for pathogenesis changes (i.e. *In vitro* changes in the binding affinity of spike protein receptor binding domain to the ACE2 receptor), changes in neutralization activity of anti-SARS-CoV-2 antibodies, and gene detection drop-outs like S-gene target failure (SGTF) in diagnostic assays.

Animal studies were excluded. Studies that only reported predictive modelling data were excluded. Reviews, overviews, and news articles that presented no original data were excluded, but checked for references to primary studies, which were screened for inclusion. Title/abstract screening was completed by a single reviewer in Covidence; full text screening was completed by one reviewer and verified by a second. The data extraction form was designed in consultation with infectious disease specialists and microbiologists; data were extracted by one reviewer and verified by a second. Critical appraisal was not conducted, but will be a feature of future reports. The final report was reviewed by infectious disease experts and microbiologists engaged as content experts on our team.

## Results

The search retrieved 1796 electronic database records and 67 grey literature records, of which 16 and 7 were included, respectively (see Appendix B: PRISMA Flow Diagram). Of the 23 sources identified, 13 were pre-prints, three were published in peer reviewed journals, and seven were grey literatures sources. Three sources report on P.1^6,12,13^, three sources report on B.1.351^13–15^ and 20 sources report on B.1.1.7^4,13,15–32^ (Table 1). One source reported on both the SA VOC and UK VOC^15^ and one reported on all three.^13^ Most studies (n=13) report on data from the UK, three studies reported on data from the United States^20,23,25^, two from Brazil,^6,12^ and one each from Israel,^17^ Wales,^32^ and Zambia.^14^ Two studies reported on data from multiple countries.^13,15^

**Table 1.** Summary of Sources

| Author | Date | Source | Objective | Sampling & Outcome of interest | Variant | Country | Main finding relevant to transmission |
| --- | --- | --- | --- | --- | --- | --- | --- |
| **WALKER ET AL<sup>16</sup>** | Jan 15, 2021 | medRxiv | To investigate whether B.1.1.7 variant is associated with higher infection rates, overall or particularly in children. | RT-PCR nasal and throat swabs from Sept 28, 2020-January 2, 2021 from 372,626 participants across the UK. RT-PCR tested for N protein, S protein, and ORF1ab. Positives for B.1.1.7 must have included N protein and ORF1ab (and a negative 2-gene for SGTF). Cycle threshold (Ct) values considered a proxy for viral load. | B.1.1.7 | UK | Identified an increase in SGTF positivity (B.1.1.7) while non-SGTF growth was stable or decreasing, with higher infection rates in adults and children. Found no evidence that Ct values were intrinsically substantially lower in SGTF-positives. |
| **MUNITZ ET AL<sup>17</sup>** | Feb 19, 2021 | medRxiv | To explore the transmission dynamics of B.1.1.7 variant and to analyze the effect of three noteworthy programs namely, mass PCR testing, focused protection of the elderly and an unparalleled prioritized vaccination program. | Analyzed >300,000 RT-PCR general community and nursing home sample between December 6, 2020 to February 10, 2021. Reported on the effective reproduction number (Rt), defined as the expected number of new infections at time t caused by an infectious individual. | B.1.1.7 | Israel | Within a six-week period B.1.1.7 became the dominant strain compared to wildtype SARS-CoV-2 (more than 90% of positive test). B.1.1.7 was also 45% more transmissible than the wildtype stain. The rise was seen more prominently in population aged 0-59. This finding attributed to the vaccine program targeting 60+ |
| **DAVIES ET AL<sup>4</sup>** | Feb 7, 2021 | medRxiv | To determine the relative transmissibility of B.1.1.7 | Over 150,000 SARS-CoV-2 samples from across the UK, as part of the COG-UK dataset as of January 2021. Growth rate of B.1.1.7 was performed using multinomial and logistical regression. | B.1.1.7 | UK | B.1.1.7 is 43-82% (CI 38-106%) higher than preexisting variants of SARS-CoV-2 in England. |
| **KIDD ET AL<sup>18</sup>** | Feb 13, 2021 | Journal of Infectious Diseases | To find evidence of B.1.1.7 variant in routine testing results in UK | RT-qPCR cycle threshold (Ct) values from 641 SARS-CoV-2 positive results between Oct 25-Nov 25, 2020. Raw Ct values were analyzed for the | B.1.1.7 | UK | Identified significantly higher proportion of SGTF positive samples compared with other gene targets ORF or N gene (27% vs 2%; |
| | | | | presence of ORF1ab, N, or S-gene target. No signal detected were defined as SGTF. Viral loads were based on lab performance for the Qnostics EQA panel. | | | p<.000001). Significantly more SGTF had higher inferred viral loads between $1 \times 10^7$ and $1 \times 10^8$ |
| **LEUNG ET AL<sup>19</sup>** | Jan 7, 2021 | Euro Surveillanc e | To compare the transmissibility of B.1.1.7 to the 501Y Variant 1 | Comparative transmissibility based on 7,003 genome sequences of SARS-CoV-2 from the GISAID database on December 19, 2020. | B.1.1.7 | UK | Findings indicate that B.1.1.7 is estimated to present an $R_0$ 1.75 times higher than 501N, meaning it is 75% more transmissible compared with the 501N strain. |
| **LIU ET AL<sup>20</sup>** | Feb 5, 2021 | bioRxiv | To test whether B.1.1.7, 20I/501Y.V1 is more transmissible due to increased bonding affinity. | Binding affinity of VOC RBD for ACE2, using a sample of 293F cells. | B.1.1.7 | US | B.1.1.7 variant determined to be much more contagious than the original version (N501-RBD). This mutated version of RBD was found to bind to human ACE2 ~10 times more tightly than the native version (N501-RBD). |
| **GRAHAM ET AL<sup>21</sup>** | Jan 29, 2021 | bioRxiv | To examine the association between the regional proportion of B.1.1.7 and reported symptoms, disease course, rates of reinfection and transmissibility | Self-reported results of 36,920 COVID Symptom Study app participants, between 28 September and 27 December 2020. Association between regional proportion of B.1.1.7 and participant symptoms, demographics, disease course, rates of reinfection and transmissibility. | B.1.1.7 | UK | An increase reproduction number $R(t)$ 1.38 (95% CI 1.06-1.71) associated with B.1.1.7 variant was found. No associations found between the proportion of B.1.1.7 in circulation and disease severity, hospitalizations, or the proportion of individuals with long symptom duration. No evidence of increased rates of infection with B.1.1.7. |
| **HUNTER ET AL<sup>22</sup>** | Jan 4, 2021 | medRxiv | To determine what value the national lockdown had on controlling the epidemic in England and particularly to investigate the impact of this pre-lockdown surge in infections. | Daily case data from 315 local authorities, available through the English Department of Health and Social Care daily COVID dashboard. Incidence and R-value per jurisdiction. | B.1.1.7 | UK | Following the lockdown there was a mean reduction in R value of 0.7 across all locals. The new variant was estimated to add .4 or greater to the R value across all locals. |
| **KISSLER ET AL<sup>23</sup>** | Feb 19, 2021 | medRxiv | To test whether acute infection of B.1.1.7 is associated with higher concentration/ longer duration of viral concentration. | Assessed longitudinal PCR tests performed in a cohort of 65 individuals affiliated with a professional sport league infected with SARS-CoV-2 undergoing daily surveillance testing, including seven infected with B.1.1.7, as confirmed by whole genome sequencing. | B.1.1.7 | US | SARS-CoV-2 variant B.1.1.7 may cause longer infections (13.3 days vs 8.2 days) with similar peak viral (Ct 19.0 vs 20.2) concentration compared to non-B.1.1.7 SARS-CoV-2, and this extended duration may contribute to B.1.1.7 SARS-CoV-2's increased transmissibility. |
| **GOLUBCHIK ET AL<sup>24</sup>** | Jan 15, 2021 | medRxiv | To determine if new variant results in higher viral loads. | All available SARS-CoV-2 sequence data and associated metadata from across the UK, as part of the COG-UK dataset as of January 2021.<br>Sequenced RT-QPCR SARS-CoV-2 positive samples. Used log<sub>10</sub> (mapped reads) as a proxy for viral load. Also reported Ct values. | B.1.1.7 | UK | Across all data there was a 3-fold higher (sequence derived) viral load in the 501Y variant compared with 501N variant (p=0.014). Differences in viral load were identified between geographic locations. Samples from Greater London had significantly lower viral loads than other locations (p=0.0016) and the association between Y501 and higher viral loads was not significant in this region (p=0.91) |
| **WASHINGTON ET AL<sup>25</sup>** | Feb 7, 2021 | medRxiv | To investigate the prevalence and growth dynamics of the variant in the United States. | Sequenced 212 B.1.1.7 SARS-CoV-2 genomes collected from testing facilities in the U.S. from December 2020 to January 2021.<br>Used RT-qPCR testing anomaly of SGTF as proxy for variant detection. Proportion of variant in daily positive cases | B.1.1.7 | US | As of the date of publication the B.1.1.7 growth rate has increased by 35-45% across the country, doubling in relative frequency about every week and a half. |
| **ZHAO ET AL<sup>26</sup>** | Jan 28, 2021 | Journal of Travel | To develop a framework to infer increased | SARS-CoV-2 sequencing data and COVID-19 surveillance data was collected from 1 | B.1.1.7 | UK | 501Y variant is estimated at 52% (95% CI: 46-58) more transmissible |
|  |  | Medicine | transmission with new variant (later identified as B.1.1.7) | August to 31 December 2020, via the public databases released by GISAID and WHO. Number of cases, reproduction number, proportion of cases with 501Y variant. |  |  | than the 501N variant, which indicates a transmission advantage of N501Y substitution. |
| **PUBLIC HEALTH ENGLAND**<sup>27</sup> | Dec 15, 2020 | Unknown | To determine prevalence and growth rates of B.1.1.7 variant | All genome sequence data available from the Milton Keynes Lighthouse Laboratory as of December 2020 were assessed and prevalence of variant per positive cases in geographic region reported. | B.1.1.7 | UK | Indication that B.1.1.7 grows 71% (95% CI: 67%-75%) faster per generation (6.5 days), yet consistent frequency does not indicate a constant selective advantage of B.1.1.7 |
| **PUBLIC HEALTH ENGLAND**<sup>28</sup> | Dec 21, 2020 | Unknown | To provide an update on the SARS-CoV-2 Variant of Concern 202012/01 | 962 genome sequences from Kent, UK, available through the COG-UK database. Prevalence and transmission rate. | B.1.1.7 | UK | The VOC (B.1.1.7) has grown rapidly in the UK and has been assessed as having substantially increased transmissibility. |
| **COLLIER ET AL**<sup>29</sup> | 2020 | medRxiv | To assess immune response following vaccination with mRNA-based vaccine BNT162b2 | Neutralizing antibody responses of 23 adults who had received the BioNTech vaccine using pseudo viruses expressing the wild-type Spike protein or the 8 mutations found in the B.1.1.7 Spike protein | B.1.1.7 | UK | Enhanced binding of the B.1.1.7 receptor binding domain to human ACE2 might be associated with ongoing transmission and possibly reduced opportunity for antibody binding. |
| **NERVTAG**<sup>30</sup> | Dec 18, 2020 | Unknown | To review the risk of transmission of VUI-202012/01 | Transmissibility based on four analytic approaches: growth rate, R-value correlation, PCR ct values, and viral load from unique genome reads from multiple databases across the UK. | B.1.1.7 | UK | Moderate confidence that B.1.1.7 demonstrates a substantial increase in transmissibility compared to other variants. |
| **NERVTAG**<sup>31</sup> | Jan 21, 2021 | Unknown | To report updated and additional analyses which strengthen the previous finding of increased disease severity in people infected with VOC B.1.1.7 | Risk of hospitalization and death based on assessments from multiple databases across the UK. | B.1.1.7 | UK | It is likely that infection with VOC B.1.1.7 is associated with an increased risk of hospitalization and death compared to infection with non-VOC viruses |
| **WELSH GOVERNMENT**<sup>32</sup> | Jan 7, 2021 | Unknown | To provide an update on the VOC 202012/01 for Wales | All available S-gene data for Wales as of 2nd January 2021.<br>Risk of transmission based on R-value and Rt | B.1.1.7 | Wales | The new VOC 202012/01 is more transmissible than the current 'wild type' SARS CoV-2, and is likely to become the dominant variant in Wales across the whole population in a matter of weeks |
| **CDC**<sup>14</sup> | Feb 26, 2021 | Unknown | To provide an update on the detection of B.1.351 VOC in Zambia | Analysis of 23 SARS-CoV-2 specimens collected between 16th to 23rd December from the University of Zambia and PATH Prevalence and viral load | B.1.351 | Zambia | Spread of the B.1.351 variant is of public health concern because of the potential for increased transmissibility and thus, increases in cases, hospitalizations and deaths. |
| **NAVECA ET AL**<sup>6</sup> | 2021 | Research Square | To determine the genetic epidemiology study of emergence of P.1 variant of concern | Generated 250 SARS-CoV-2 high-quality whole-genome sequences from individuals living in 25 municipalities of the Amazonas state between 16th March 2020 and 13th January 2021 | P.1 | Brazil | P.1 prevalence in the Amazonas state increased from 0% in November 2020 to 73% in 1st-15th January 2021. There was also a significant (Mann-Whitney test, $P < 0.0001$) lower median score in URT samples from P.1-infected (Ct = 19.8) compared with non-P.1-infected (Ct = 23.0) subjects, indicating that viral load was ~ 10-fold higher in P.1 infections than in non-P.1 infections. |
| **FARIA ET AL**<sup>12</sup> | Jan 12, 2021 | medRxiv | To report on the evolution and emergence of the P.1 variant of concern | Analysis of 436 SARS-CoV-2 RT-qPCR positive results from a testing laboratory in Manaus from 1 November 2020 to 9 January 2021.<br>Molecular clock analysis, transmissibility and immune evasion | P.1 | Brazil | It cannot yet be determined if P.1 infection is associated with increased viral loads or a longer duration of infection. |
| **GRABOWSKI**<br>**ET AL<sup>15</sup>** | Feb 21,<br>2021 | medRxiv | To determine the growth ratio of VOC between Oct 19-Dec 20 and the estimated replicative advantage of B.1.1.7 and B.1.351 compared to 20A.EU1 strain. | SGTF as proxy for prevalence of the VOC genome. Aggregated weekly data from England and South Africa submitted to GISAID and samples collected in Wales, Scotland, Denmark and US. Relative replication estimate, assuming the same average serial interval of 6.73 days | B.1.1.7, B.1.351 | UK, Wales, Denmark, Scotland, US | The replicable advantage of the B.1.1.7 over the A20.EU1 strain is in the range of 1.83-2.18 in the UK and 1.69-1.76 in Denmark, Scotland, Wales and the US. The replicable advantage of the B.1.351 was estimated at 1.55. |
| **WHO<sup>13</sup>** | Feb 23,<br>2021 | Unknown | To provide an update on ongoing studies and the geographical distribution of select VOCs as reported by countries as of 23 February 2021. | Analysis of official laboratory-confirmed COVID-19 cases and deaths as reported to WHO by countries, territories and regions. Global prevalence and emerging information on key VOCs | B.1.1.7, B.1.351, P.1 | International | Increased transmissibility has been reported for some VOCs resulting in increased incidence in settings experiencing community transmission. |

**Table 2.** Summary of Transmissibility

| **VOC** | **Author, Year** | **R value (R<sub>t</sub> and R<sub>0</sub>)\*** | **Region** |
| --- | --- | --- | --- |
| **B.1.1.7** | Munitz, 2021 | R<sub>t</sub>: 1.71 (95% CI: 1.59-1.85) | Israel |
|  | Public Health England, 2020 | R<sub>t</sub>: 1.47 (95% CI: 1.34-1.59) | Kent, UK |
|  | Graham, 2021 | R<sub>t</sub>: 1.35 (95% CI: 1.02-1.69) | Scotland, Wales and 7 NHS regions in England |
|  | Zhao, 2021 | R<sub>t</sub>: 52% (95% CI: 46, 58) | GISAID database (International) |
|  | Davies, 2021 | R<sub>0</sub>: 78% (95% CI: 69-87%) | UK |
|  | Leung, 2021 | R<sub>0</sub>: 75% (95% CI: 70-80%) | UK (Fall 2020) |
|  | Hunter, 2021 | A) R<sub>t</sub>: 1.3 (95% CI: NR)<br>B) R<sub>t</sub>: 1.3 (95% CI: NR)<br>C) R<sub>t</sub>: 1.1 (95% CI: NR) | A) East of England<br>B) London<br>C) South East England |
|  | Grabowski, 2021 | R<sub>t</sub>: 1.83-2.18 (95% CI: 1.71-2.40) | GISAID database (international) |
| **B.1.351** | Grabowski, 2021 | R<sub>t</sub>: 1.55 (95% CI: 1.43-1.69) | GISAID database (international) |
| **P.1** | NR | NR | NR |

\*values presented as reported by study authors

### Limitations

As this was a rapid scoping review, **formal critical appraisal of included studies was omitted**. The next update of this report will include critical appraisal of this and other literature identified in the updated search. Of note, most included sources have not been peer-reviewed. Considering this and the fact that this is an emerging field of research, findings should be interpreted with caution until more evidence becomes available. We also identified significant heterogeneity between studies in terms of analysis and reporting, testing approaches for the VOCs, and sample size. A separate search for VOC definitions was not executed due to time constraints. Data informing question 3 was extracted from studies included for question 1 and 2.

### Question 1: How much more transmissible are the VOCs?

To address question 1, we searched for data related to risk of transmission, changes in transmission, and reproduction number. We describe the findings by each VOC. When interpreting these findings, it is important to consider that transmission can be reflected in a number of ways, including reproduction number, VOC growth rate, and gene detection drop-outs like SGTF. Each measure used has limitations and potential biases depending on how it was used, and the population assessed.

#### B.1.1.7

According to the WHO as of March 9^th^, 2021, B.1.1.7 has been reported in 111 countries across all six WHO regions.^8^

In terms of population transmission risk, Munitz et al. reported that prior to December 22^nd^, 2020 in Israel, B.1.1.7 was undetectable within a pool of positive cases, but within 4 weeks, the B.1.1.7 became the dominant strain (by January 21^st^), reaching to 92% following 2 more weeks.^17^ They also reported that B.1.1.7 was 45% more transmissible than the wild type in Israel based on an analysis of over 300,000 RT-PCR tests from the general community and a nursing home sample between December 6^th^, 2020 and February 10^th^, 2021. There was no evidence of variation in risk of transmission based on age, with the exception of a population of individuals >60 years, approximately 2 weeks post-first vaccination, which showed a decline. Munitz et al. found that prior to the impact of the national lockdown in Israel, the Rt of B.1.1.7 was as high as 1.71 (95% CI:1.59-185) compared to 1.12 (95% CI: 1.10-1.15) for the wild type.^17^

In the UK, Zhao et al. estimated that B.1.1.7 is 52% (95% CI: 46, 58) more transmissible than the wild type based on 94,934 SARS-CoV-2 sequencing data and COVID-19 surveillance data from August 1^st^ to December 31^st^, 2020 as available in the public databases released by GISAID and World Health Organization.^26,28^

In a December 18^th^, 2020 NERVTAG report, they found that the B.1.1.7 VOC growth rate was 71% (95% CI: 65%-75%) higher than other variants.^30^ Similarly, the Public Health England December 21^st^, 2020 report found the growth factors as 1.47 (95% CI: 1.34-1.59) between October 25^th^ and December 5^th^, 2020 using SGTF among PCR positives as a proxy for B.1.1.7.^28^

According to Public Health England, B.1.1.7 VOC was found to be significantly associated with higher reproduction number.^28^ Rt increased by 0.57 (95% CI: 0.25-1.25) in fixed model, while a random effect model gave an additive effect of 0.74 (95%CI: 0.44-1.29) based on 1419 VOC genomes and 33,792 non-VOC genomes. Adjusting rates of SGTF for variable specificity over time and between local authorities, the same models estimated an increase of Rt of 0.52 (95%CI: 0.39-0.70) in a fixed effect model while a random effect model gave an estimated additive effect of 0.60 (95%CI: 0.48 - 0.73). Similar estimates of 0.56 (95%CrI: 0.37-0.75) were obtained using a Bayesian regression model accounting for errors in VOC frequencies and Rt estimates.

In the United States, Washington et al. assessed SGTF as a proxy to B.1.1.7 from December 2020 to January 2021. They calculated that B.1.1.7 had an increased transmissibility of 35-46% across the US, with rates of 29-37% in California and 38-49% in Florida.^25^

Walker et al. reported that the growth rate in SGTF positives (proxy for B.1.1.7) generally exceeded the decline in non-SGTF SARS-CoV-2 positive cases in the UK, by an average of 6%, suggesting an overall increase in rate of VOC infections, not just replacement.^16^ There was no evidence of variation based on age.

Grabowski et al. reported that B.1.1.7 started expanding in the UK during week 43 (<5 cases were reported per week) and by week 51, reached 57% of all sequenced genomes based on GISAID data collected between week 43 and week 51 of 2020 (October 19^th^ to December 20^th^, 2020).^15^ This was compared to the proportion of 20A.EU1 strain, characterized by A222V substitution in spike protein which constituted more than 65% of genomes sequenced in England in November 2020, which dropped from 86% to 35% and other genomes dropped from 31% to 8%.^15^ Grabowski et al. found the Rt for the B.1.1.7 over 20E.EU1 was between 1.83-2.18 (95% CI: 1.71-2.40) between weeks 43 to 51 (8 weeks) in the UK.^15^ The Rt for B.1.1.7 to other strains was calculated to be between 2.03-2.47 (95% CI: 1.89-2.77), where the lower bound is the estimate between weeks 44-51 and the higher bounds for weeks 43-47 in the UK.

Graham et al. calculated the relative advantage of B.1.1.7 to the 20A.EU1 strain, between weeks 43-51 in the UK, which was calculated as 1.96-fold per week.^21^ For the relative advantage of the B.1.1.7 VOC to 20A.EU1 strain, it was calculated as 1.65 for Wales (95%CI: 1.56-1.83), 1.73 for Denmark (95%CI: 1.64-181), 1.71 for Scotland (95%CI: 1.52-1.93), and 1.76 for the US (95%CI: 1.48-2.09). In all four countries calculated, the VOC strain was less prevalent than it was in England. Graham et al. also found that the mean of the additive increase in R for B.1.1.7 was 0.34 (95% CI: 0.02-0.66), and the multiplicative increase was 1.35 (95% CI 1.02-1.69).^21^ Using the SGTF data, with analysis limited to the period after 1 December when at least 95% of all SGTF cases were B.1.1.7, the Rt of B.1.1.7 has an additive increase of 0.26 (0.15-0.37) and a multiplicative increase of 1.25 (1.17-1.34).

Davies et al. found that the B.1.1.7 was calculated to increase the basic reproduction number R_0_ by 78% (95% CI: 69-87%) in England.^4^ Leung et al. found that the R0 of B.1.1.7 was 75% (95% CI: 70– 80%) higher than that of the wildtype strain (501N) in a UK sample.^19^

Hunter et al. found that from mid-lockdown in November to the first week in December, most variants declined but the new variant increased quite dramatically during that same time.^22^ The estimated R_t_ value for the East of England, London and South East regions for the novel variant was 1.3, 1.3 and 1.1 respectively indicating growth in the epidemic of new variant cases. The equivalent R_t_ values for all other strains combined were 0.92, 0.91 and 0.97.

No data was found on greater susceptibility to infection given exposure or in greater infectiousness once infected for the B.1.1.7.

### SUMMARY

<u>The majority of information available on transmission related to VOCs is on the B.1.1.7 variant. Based on limited research, B.1.1.7 VOC transmission risk ranges from 45-71% higher. The R_0_ of B.1.1.7 compared to non-VOC ranges from 75-78% higher, while the reported R</u>_t_ <u>values range from 1.1-2.8. Additionally, it appears there is an additive transmission effect, where it is not only replacing previous infections, but also associated with an increase in the number of infections. However, further work is needed to clarify and expand on this, recognizing the possibilities of inherent biases in study design or data captured.</u>

#### B.1.351

According to the WHO as of March 9^th^, 2021, B.1.351 has been reported in 58 countries across all six WHO regions.^8^

In terms of transmission risk, Grabowski et al. reported that the Rt for B.1.351 was 1.55 (95%CI: 1.43-1.69), meaning that it is 55% more transmissible than the wild type strain.^15^ They also reported that the relative advantage ratio of B.1.351 to other strains was at a weekly rate of 1.58 (95%CI: 1.45-1.72). The CDC reported that the detection of the B.1.351 variant coincided with a rapid rise in confirmed cases in Zambia, establishing an epidemiologic linkage between COVID-19 outbreaks in Zambia and South Africa.^14^ However, they did not report more specifics on the rate of transmission.^13^ While the WHO report stated that the B.1.351 variant was more transmissible, this was not supported by their references as Davies et al.^4^ reports on B.1.1.7 and Wibmer et al.^33^ does not seem to report on transmissibility.

While there have been no reported findings related to greater susceptibility to infection given exposure to B.1.351 nor in greater infectiousness once infected, the WHO report cited a case study that found a person was re-infected with the B.1.351 strain after recovery from an earlier COVID-19 infection and the clinical presentation was more severe.^34^ As this was a letter to the editor case study that did not report on transmission beyond a single case, it did not meet our eligibility criteria to include as an original source.

### SUMMARY

<u>While B.1.351 might be more transmissible based on findings from one study in the UK, limited recommendations or conclusions can be drawn at this time.</u>

#### P.1

According to the WHO as of March 9^th^, 2021, P.1 has been reported in 32 countries across all six WHO regions.^8^

Faria et al. used dynamic modelling that integrates genomic and mortality data, to estimate that P.1 may be 1.4 to 2.2 times more transmissible and 25-62% more likely to evade protective immunity of previous infections on non-VOCs.

Naveca et al. reported P.1 was first detected on December 4^th^, 2020 in Manaus, Brazil and displayed an extremely rapid increase in prevalence up to January 2021.^6^ In particular, through sequencing all SARS-CoV-2 positive samples between November 1^st^, 2020 and January 31^st^, 2021, they found that none of the samples genotyped by real-time PCR before December 16^th^ had the NSP6 deletion, indicating a very low prevalence of P.1 before mid-December 2020 in Amazonas. Using both genomic sequencing and real-time PCR testing, P.1 prevalence in the Amazonas state increased from 0% in November 2020 (n = 0/88), to 4% in December 1^st^ to 16^th^, 2020 (n = 2/54), 45% in December 17^th^ to 31^st^, 2020 (n = 104/232), and 73% in January 1^st^ to 15^th^, 2021 (n = 119/162).

No data was found on greater susceptibility to infection given exposure or on greater infectiousness once infected for the P.1 VOC.

### SUMMARY

<u>While P.1 might be more transmissible based on findings from two studies in Brazil, limited recommendations or conclusions can be drawn at this time due to a paucity of data.</u>

### Question 2: Why are they more transmissible?

To address question 2, we searched for data related to the mechanism of transmission (e.g., peak viral load, transmission in relation to exposure). It is worth noting that many studies assessed viral load through analysis of RT-qPCR Ct values from laboratory testing of respiratory samples. Ct values are only a proxy measure of peak viral load. Ct measure can vary based on a number of factors including instrument, type of specimen (e.g upper or lower respiratory tract), timing of sample related to onset of symptoms or exposure, and storage or transport of specimen.^30,35–37^

#### B.1.1.7

Several studies have focused on measuring viral load as an indicator of increased transmissibility of B.1.1.7. In general, we identified variation across studies in terms of methods and outcomes. A large surveillance study conducted in the UK between September 28^th^, 2020 and January 2^nd^, 2021, with monthly PCR tests of nose and throat swabs on children 2-15 years old (n=372,626) identified an increase in SGTF, including during periods of lockdown when non-SGTF strains were stable or decreasing.^16^ However, there was no evidence that viral loads were lower in those who were SGTF positive and there was no difference in growth between children and adults counter to other studies which suggest B.1.1.7 is particularly adapted to transmit more in children. Further, there was no evidence to suggest that increased infection rates were mediated by higher viral loads (lower Ct).^16^ To assess the surveillance testing program in nursing homes in Israel, Munitz et al. analyzed primary data from RT-PCR samples collected between December 6^th^, 2020 and February 10^th^, 2021 to compare Ct threshold values of 60+ age groups in the general community and nursing homes. Findings revealed significantly lower Ct values in the community in comparison to nursing homes residents with matching age groups (p,0.001).^17^ These results were deemed to support the success of the surveillance program as viral load was understood to drive transmission. Kidd et al. analyzed a dataset of 641 SARS-CoV-2 positive results of respiratory samples from October 25^th^ to November 25^th^, 2020 and identified a significantly higher number of SGTF samples were associated with a higher viral load, although similar viral loads were seen in non-SGTF samples.^18^

Golubchik et al. sequenced RT-QPCR CoV-2 positive samples from four UK Lighthouse Laboratories and identified 88 SARS-CoV-2 positive samples with the spike protein N501Y amino acid substitution (proxy of any of the 3 circulating VOCs) taken between October 31^st^, 2020 and November 13^th^, 2020. They identified approximately 3-fold higher median viral load in the 501Y containing samples compared with wildtype N501 samples. Results remained significant when controlled for batch but not Lighthouse laboratory.^24^

The December 18^th^ report by NERVTAG concluded that there were insufficient data to draw any conclusions on underlying mechanisms of increased transmissibility, including increased viral load.^30^

As the number of B.1.1.7 infections have grown rapidly, studies have also examined if the location of the 501Y mutation within the spike protein receptor binding domain (RBD) and increased affinity for the ACE2 receptor might provide a biological hypothesis for greater infectivity. Liu et al. tested the affinity of B.1.1.7.201/501Y.V1 for ACE2 receptor and found that the mutated spike protein RBD with a N501Y mutation had a ∼10 times greater affinity for ACE2 than the wild-type N501-RBD *in vitro*. This provides a possible hypothesis for potentially explaining the increased rate of infections in the United Kingdom.^20^

Studies have also examined how changes in social contact patterns correlate with changes in transmission of B.1.1.7. Davies et al. used Google mobility and social contact survey data from September 2020 to December 2020 to examine the influence of social contact patterns on transmission. A national lockdown was imposed between November 5^th^, 2020 and December 2^nd^, 2020, however, no differences in social interactions were identified between regions of high and low B.1.1.7 prevalence. The authors suggest the lack of correlation between social contact patterns and transmission may be related to evolution of the virus.^4^

The length of infection has also been examined as a means for understanding transmissibility of B.1.1.7 When compared with non-VOC, B.1.1.7 is reported to have a longer period of detectability by RT-PCR (13.3 days [CrI: 10.1, 16.5] compared with 8.2 days [CrI: 6.5,9.7]) with similar peak viral concentration (19 Ct [15.8,22] versus 20.2 Ct [19.1,21.4]) suggesting greater chance of transmission with B.1.1.7 over a long period of time.^23^

#### B.1.351

WHO reports there is growing evidence that mutations present in B.1.351 may help the virus evade immune system responses triggered by previous infections of SARS-CoV-2.^13^

The CDC reports that B.1.351 might be associated with higher viral loads and contain a spike protein that might hinder antibody binding which could lessen naturally developed immunity, but this was not substantiated by primary data included in this review.^14^

#### P.1

Faria et al. noted that similar to other VOCs, P.1 is associated with increased binding to the human ACE2 receptor, but they were unable to determine if P.1 infection was associated with increased viral loads or longer infection.^12^

Naveca et al. used real-time RT-PCR Ct values as a proxy of viral load and compared samples of P.1 positive and P.1 negative collected at the similar time from onset of symptoms. Analysis revealed a significantly (p<0.0001) lower median score in P.1 positive (Ct=19.8) compared with P.1 negative (Ct=23.8) indicating approximately 10-fold higher viral load in P.1 infections, suggesting that P.1 infected adult individuals are more infectious than those with non-P.1 viruses. These findings hold true when comparing Ct values for adult (18-59 years) men (P=0.005), adult (18-59 years) women (p<0.0001) and older (>59 years) women (p=0.0149) but not in older (>59 years) men (p=0.4624).^6^

### Summary

<u>While it is difficult to draw conclusions about the underlying mechanisms of VOC transmissibility from the existing data, there is some evidence to suggest there may be an increase in viral load using RT-PCR Ct values as a proxy measure. A few studies have also proposed that VOCs might evade immune system response through increased ACE2 binding. We identified variation in methods, outcome measures and reporting across included studies and limited evidence related to the underlying mechanisms of transmissibility, particularly on P.1 and B.1.351.v</u>

### Question 3: What criteria are used to define new variants of concern?

A report from Public Health England first identified an emerging SARS-CoV-2 variant as *Variant Under Investigation (VUI 202012/01)* on December 8^th^, 2020. The nomenclature of this variant was later changed to *Variant of Concern* on December 18^th^, 2020, after an expert review and risk assessment.^28^

The following components have been used to describe a VOC in the 23 included sources:

1. Phylogenetically distinct from other SARS-CoV-2 variants^12,28^
2. Mutations of biological significance^4,12^
3. Rapid spread, dominance and/or selective advantage over other variants^4,12,28,30^

On February 25^th^, 2021, the WHO released an updated working definition for SARS-CoV-2 Variants of Interest (VOI) and Variants of Concern (VOC).^13^ A VOI is named as such if:

*“It is phenotypically changed compared to a reference isolate or has a genome with mutations that lead to amino acid changes associated with established or suspected phenotypic implications” AND “has been identified to cause community transmission/multiple COVID-19 cases/clusters or has been detected in multiple countries” OR “is otherwise assessed to be a VOI by WHO in consultation with the WHO SARS-CoV-2 Virus Evolution working group*.*”*^13^

A VOC is defined by WHO if it meets the criteria for a VOI as stated above and is associated with:

*“Increase in transmissibility or detrimental change in COVID-19 epidemiology; increase in virulence or change in clinical disease presentation; decrease in effectiveness of public health and social measures or available diagnostics, vaccines, therapeutics” OR “assessed to be a VOC by WHO in consultation with the WHO SARS-CoV-2 Virus Evolution working group*.*”*^13^

## Discussion

In this rapid scoping review, we found 23 sources that provide insight into the transmission risk of the three main SARS-CoV-2 VOCs as of March 2021 (B.1.1.7 or UK variant; B.1.351 or SA variant; P.1 or Brazil variant). The majority of information available on transmission is on the B.1.1.7 variant. Based on limited research, B.1.1.7 VOC transmission risk ranges from 45-71% higher and the R_t_ of B.1.1.7 compared to non-VOC ranges from 75-78% higher. Additionally, it appears there is an additive transmission effect, where it is not only replacing previous infections, but also associated with an increase in the number of infections. While there is heterogeneity in the data, the evidence points towards an increase in transmissibility in VOCs. Further work is needed to clarify and expand on this. While B.1.351 and P.1 might be more transmissible based on findings from one study each, limited recommendations or conclusions can be drawn at this time due to a paucity of data. Additionally, currently no data is available on greater susceptibility to infection given exposure or in greater infectiousness once infected for all the VOC, with one case study noting a possibility of re-infection for the B.1.351 VOC.

With respect to understanding why VOCs are more transmissible, it is difficult to draw conclusions from existing data on the underlying mechanisms of increased transmission due to heterogeneity of included studies and an overall dearth of evidence, particularly on P.1 and B.1.351. There appears to be a lack of consistent findings related to whether viral loads (approximated by Ct values) are higher in VOC-infected individuals, and whether the period of RT-PCR positivity may be extended, with variation depending on the population sampled, specimen type, testing method and timing. Further it is unclear if the VOC spike protein interferes with the immune response through increased ACE2 binding. *In vitro* and animal data have been excluded from this review and may contribute to findings in future reports.

While several studies and governing bodies have reported key characteristics of VOCs, there is variation among the terminology. The WHO released a working definition for both VOIs and VOCs in February 2021,^13^ but this may continue to evolve as more evidence on circulating variants emerges.

### Potential Implications

- Continued monitoring of circulating and emerging VOCs at a regional, national and international level is important. It is also important that this information is shared with national and international stakeholders and health organizations in a timely manner.
- Continued routine systematic sequencing of SARS-CoV-2 and VOCs with a focus on using a standardized testing approach to facilitate data comparison across multiple locations.
- As VOCs appear to be as transmissible if not more than the non-VOC strains, existing public health guideline remain important. Any lifting public health measures, including re-opening or “returning to normal”, should be done cautiously.
- If synergized and shared, information about public health precautions and restrictions can be included as a dimension in associative or epidemiological studies, which will be important to interpret VOC data in the local context.
- While current evidence is mixed, this may be related to differences in measurement approaches, including Ct and R as surrogate measures of viral load and transmission, respectively, across studies. Additionally, Ct measure can vary based on a number of factors including instrument, type of specimen (e.g. upper or lower respiratory tract), timing of sample related to onset of symptoms or exposure, and storage or transport of specimen. Future surveillance studies should report more details on characteristics of testing, if available.
- Further research is needed on the biological relevance of mutations observed in VOCs, as well as the pathophysiology of why transmission is higher, as the mechanism is still not well understood.

## Data Availability

All datasets supporting the conclusions of this article are included within the article.

## Funding Acknowledgement(s)

The SPOR Evidence Alliance (<u>SPOR EA</u>) is supported by the Canadian Institutes of Health Research (<u>CIHR</u>) under the Strategy for Patient-Oriented Research (<u>SPOR</u>) initiative.

COVID-19 Evidence Network to support Decision-making (<u>COVID-END</u>) is supported by the Canadian Institutes of Health Research (<u>CIHR</u>) through the Canadian 2019 Novel Coronavirus (COVID-19) Rapid Research Funding opportunity.

## Definitions

**20A.EU1**: also known as B.1.177, first identified in Spain, contains a mutation called A222V on the viral spike protein

**ACE2**: Angiotensin Converting Enzyme 2, receptor site on cells including where SARS-CoV-2 binds

**B.1.1.7**: variant of concern originating in the United Kingdom, also known as VUI 202012/01 and VOC 202012/01

**B.1.351**: variant of concern originating in South Africa

**P.1**: variant of concern originating in Brazil, also known as B.1.28.1

**CDC**: Centers for Disease Control and Prevention

**CI**: **c**onfidence interval

**COG-UK:** COVID-19 Genomics UK

**CrI**: credible interval

**GISAID**: Global Initiative on Sharing All Influenza Data

**Ct**: cycle threshold, provides a relative measure of viral quantity

**PCR**: polymerase chain reaction, method for DNA replication and genome sequencing

**NERVTAG**: New and Emerging Respiratory Virus Threats Advisory Group

**RBD**: receptor binding domain, fragment of a virus which binds to a specific receptor to gain entry to a cell

**R**_**t**_: effective reproduction number, number of new cases generated in a population at a certain time period while factoring in immunity

**R**_**0**_: basic reproduction number, expected number of cases generated by one case in a population when everyone is susceptible to infection

**SA**: South Africa

**SGTF**: spike OR S gene target failure, correlates with the increase of confirmed, sequenced variants

**UK**: United Kingdom

**US:** United States

**VOC**: variant of concern

**VOI**: variant of interest

**VUI**: variant under investigation

**WHO**: World Health Organization

## Appendix A: Literature Search Strategy

### MEDLINE

*The MEDLINE search strategy was peer reviewed by another information specialist using the PRESS checklist.

*COVID-19 search filter: CADTH https://covid.cadth.ca/literature-searching-tools/cadth-covid-19-search-strings/*

|  |  |
| --- | --- |
| 1 | (coronavirus/ or betacoronavirus/ or coronavirus infections/) and (disease outbreaks/ or epidemics/ or pandemics/) |
| 2 | (ncov* or 2019ncov or 19ncov or covid19* or covid or sars-cov-2 or sarscov-2 or sarscov2 or severe acute respiratory syndrome coronavirus 2 or severe acute respiratory syndrome corona virus 2).ti,ab,kf,nm,ot,ox,rx,px. |
| 3 | ((new or novel or "19" or "2019" or wuhan or hubei or china or chinese) adj3 (coronavirus* or corona virus* or betacoronavirus* or CoV or HCoV)).ti,ab,kf,ot. |
| 4 | ((coronavirus* or corona virus* or betacoronavirus*) adj3 (pandemic* or epidemic* or outbreak* or crisis)).ti,ab,kf,ot. |
| 5 | ((wuhan or hubei) adj5 pneumonia).ti,ab,kf,ot. |
| 6 | or/1-5 [CADTH COVID-19 filter, no date limit] |
| 7 | ((uk or united kingdom or england or english or britain or british or kent) adj3 (variant* or voc or vui)) or "b117" or "20i 501yv1" or "variant of concern 202012 01" or "voc 202012 01" or "variant under investigation in december 2020" or "variant under investigation 202012 01" or "vui 202012 01").ti,ab,kw,kf. |
| 8 | ((south africa* or sa) adj3 (variant* or voc or vui)) or "b1351" or "501v2" or "501yv2" or "20h 501yv2" or "20c 501yv2").ti,ab,kw,kf. |
| 9 | ((brazil* adj3 (variant* or voc or vui)) or "p1" or "b11281" or ((mutation* or spike*) adj3 (k417t or e484k or n501y))).ti,ab,kw,kf. |
| 10 | ((mutation* or spike*) adj3 d614g).ti,ab,kw,kf. |
| 11 | or/7-10 |
| 12 | 6 and 11 |
|  | **249 results 2021-02-23** |

### Embase

*COVID-19 search filter: CADTH adapted to Embase*.*com format; line 1 exploded*

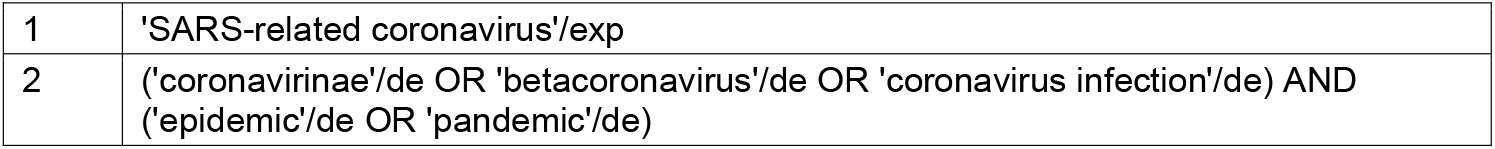
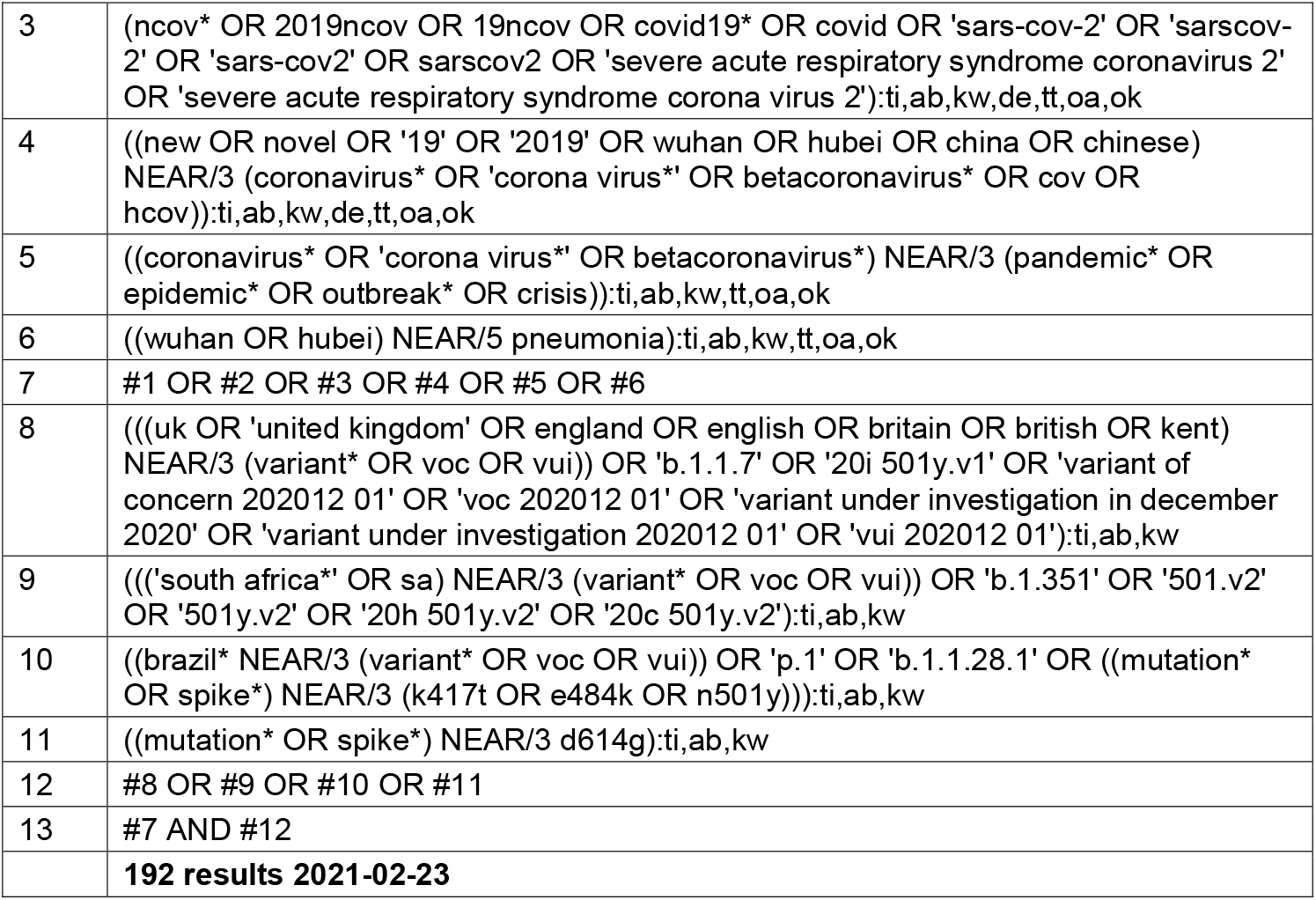

### Cochrane

|  |  |
| --- | --- |
| 1 | MeSH descriptor: [Coronavirus] this term only |
| 2 | MeSH descriptor: [Betacoronavirus] this term only |
| 3 | MeSH descriptor: [Coronavirus Infections] this term only |
| 4 | {or #1-#3} |
| 5 | MeSH descriptor: [Disease Outbreaks] this term only |
| 6 | MeSH descriptor: [Epidemics] this term only |
| 7 | MeSH descriptor: [Pandemics] this term only |
| 8 | {or #5-#7} |
| 9 | #4 and #8 |
| 10 | (ncov* or 2019ncov or 19ncov or covid19* or covid or "sars-cov-2" or "sarscov-2" or sarscov2 or "severe acute respiratory syndrome coronavirus 2" or "severe acute respiratory syndrome corona virus 2"):ti,ab,kw |
| 11 | ((new or novel or "19" or "2019" or wuhan OR hubei OR china OR chinese) near/3 (coronavirus* or "corona virus*" or betacoronavirus* or cov OR hcov)):ti,ab,kw |
| 12 | ((coronavirus* or "corona virus*" or betacoronavirus*) near/3 (pandemic* or epidemic* or outbreak* or crisis)):ti,ab,kw |
| 13 | ((wuhan or hubei) near/5 pneumonia):ti,ab,kw |
| 14 | {or #9-#13} |
| 15 | (variant* or voc or vui or mutation* or spike):ti,ab |
| 16 | #14 and #15 |
|  | **82 results in CENTRAL 2021-02-23**<br>**1 result in CDSR 2021-02-23** |

### Epistemonikos

Basic search of the following terms within the LOVE:

variant* OR voc OR vui OR “B.1.1.7” OR “20I/501Y.V1” OR “202012/01” OR “B.1.351” OR “501.V2” OR “501Y.V2” OR “20H/501Y.V2” OR “20C/501Y.V2” OR “P.1” OR “B.1.1.28.1” OR “K417T” OR “E484K” OR “N501Y” OR “D614G”

**966 results 2021-02-23**

### medRxiv / bioRxiv

medRxiv and bioRxiv simultaneous search; Date limit: October 1 2020 – present; Title and Abstract search; All words (unless otherwise specified); 50 per page; Best Match

Searches:

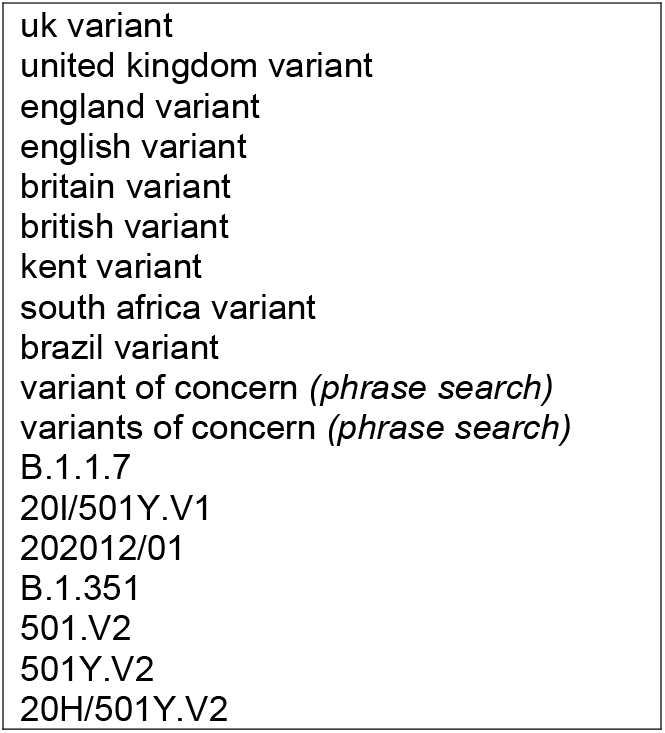
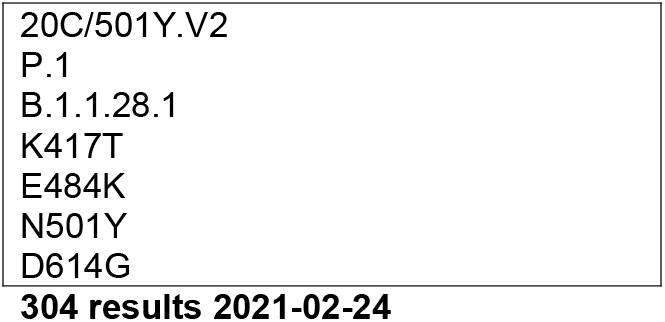

### Google

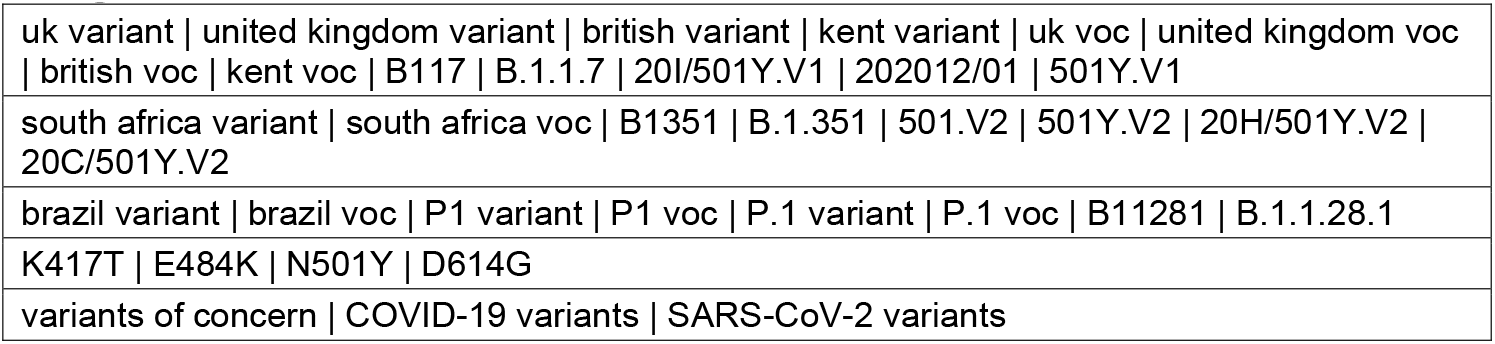

### Twitter

Basic search; Top results

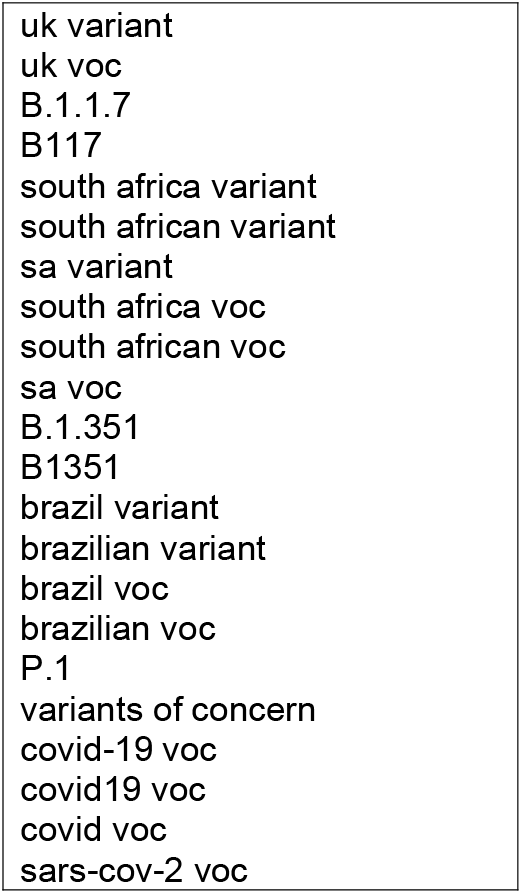

### Other websites

Manual searches of the following websites:

|  |  |
| --- | --- |
| WHO | https://www.who.int/ |
| Canadian government websites (all) – includes Health Canada and PHAC | https://www.canada.ca/en.html |
| CDC | https://www.cdc.gov/ |
| CADTH COVID-19 Evidence Portal | https://covid.cadth.ca/ |
| Public Health England | https://www.gov.uk/government/organisations/public-health-england |
| South Africa Dept of Health | http://www.health.gov.za/ |
| Brazil – Ministério da Saúde | https://www.gov.br/saude/pt-br |
| NZ Ministry of Health | https://www.health.govt.nz/ |
| Australian Dept of Health | https://www.health.gov.au/ |
| Italy - Dipartimento della Protezione Civile | http://www.protezionecivile.gov.it/ |

## Appendix B: PRISMA Diagram

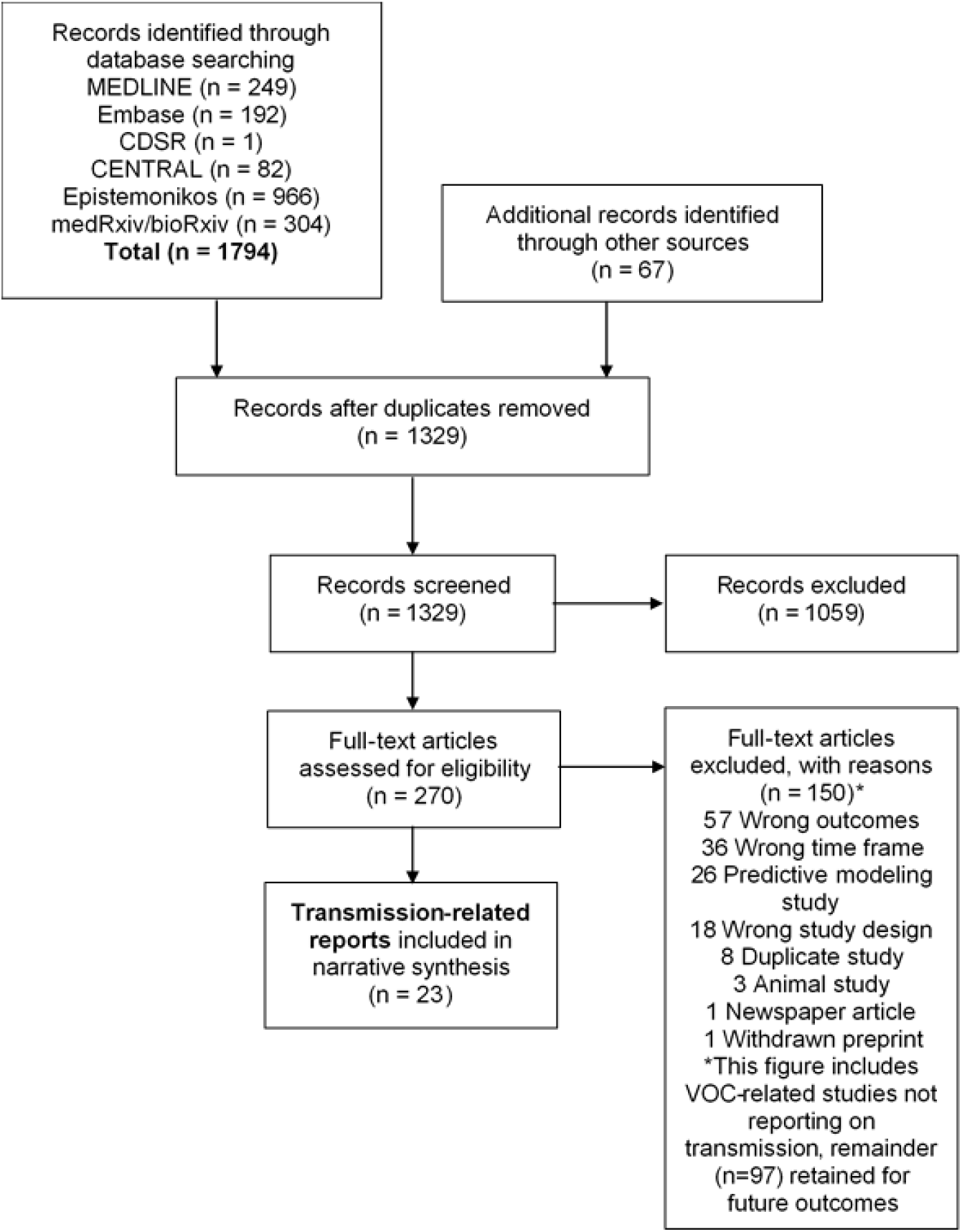

